# Recommendations for Enhancing COVID-19 Test and Treat Programs in Four African Countries: Insights and Strategies from a Qualitative Study

**DOI:** 10.1101/2025.03.24.25324159

**Authors:** Shanti Narayanasamy, Bridget C. Griffith, Sarah Gonzales, Caroline E. Boeke, Yamikani Gumulira, Nervine Hamza, Jessica T. Joseph, Allison A. Lewinski, Norman Lufesi, Alexander Martin-Odoom, Nyuma Mbewe, Elizabeth McCarthy, Carl Mhina, Julie Miller, Milimo Selena Mweemba, Faustina Ofosua Mintah, Lawrence Ofori-Boadu, Rodrigue Albert Ndayishimiye, Sean Regan, Edson Rwagasore, Krishna Udayakumar, Cameron R. Wolfe, Megan M. Oakes, Hayden B. Bosworth, the COVID-19 QuickStart Consortium

**Author notes:** These authors contributed equally to this work (Shanti Narayanasamy, Bridget C. Griffith). These authors also contributed equally to this work.

## Abstract

Sub-Saharan Africa had a significant burden of infections and deaths from COVID-19. However, throughout the pandemic, the region experienced delays and more limited access to diagnostics and treatment than high-income countries. From late 2022 to 2023, nirmatrelvir/ritonavir was introduced in four sub-Saharan African countries (Ghana, Malawi, Rwanda and Zambia) in a test and treat (T&T) model. This manuscript aims to understand the perspectives of key stakeholders, Ministry of Health, public sector personnel and health care workers on the recommendations for improvement and strengthening of national COVID-19 T&T programs deployed in the four study countries. We conducted in-depth, semi-structured interviews with individuals from two key stakeholder groups: Ministry of Health or public sector personnel involved in developing and/or implementation of COVID-19 T&T policy (purposive sampling) and healthcare workers involved in administering COVID-19 testing and/or treatment at study sites (convenience sampling). Sample size was driven by information power; our target sample size was ten to 12 interviews per stakeholder group per country. We conducted a descriptive qualitative study to explore recommendations for improvement and strengthening of national COVID-19 T&T programs using the Consolidated Framework for Implementation Research to guide qualitative data collection and analysis. Four key themes were identified by key stakeholders as critical to scaling up COVID-19 T&T programs across all study countries: increasing community education, engagement and awareness of COVID-19 and the T&T program; adjusting the T&T program to ensure program integration, decentralization and sustainability; expansion of SARS-COV-2 testing and ensuring availability of testing kits and oral antivirals through reliable national supply chains; and ensuring ongoing training and support for healthcare workers on the COVID-19 T&T program. Our finding, that recommendations were largely common across countries, suggest that a unifying framework for introducing new drugs through T&T programs during future pandemics in Sub-Saharan countries may be possible, particularly when implementation strategies can be customized to fit local contexts.

## INTRODUCTION

The COVID-19 pandemic has led to over 776 million cases globally as of December 2024, 84.8 million of which occurred in low- and middle-income countries (LMICs).^1^ As of December 2024, Sub-Saharan Africa recorded 9.6 million cases and 176 thousand deaths.^1^ While effective diagnostics and treatments have aided in reducing the number of cases and deaths from the disease globally, LMICs experienced delays and more limited access to diagnostics and treatment.^2^

Nirmatrelvir/ritonavir is an oral antiviral for the treatment of non-severe COVID-19 disease. It reduces mortality and decreases hospitalizations in unvaccinated, non-hospitalized adult patients at high risk of progression to severe COVID-19 disease.^3^ The drug was made available in high-income countries, such as the United States, from December 2021, however the first reported delivery of drug to a LMIC did not occur until March 2023 (Cambodia) after the major COVID-19 waves had passed.^4,5^

Clinical guidelines recommend that individuals begin treatment with nirmatrelvir/ritonavir as soon after symptom onset as possible, preferably within five days, to reduce the risk of progression to severe COVID-19 disease and death.^6^ Initiating treatment for COVID-19 within five-days of symptom onset is a challenge in the sub-Saharan African setting, where it was estimated in 2021 that only six in seven cases in the region were being detected.^7^ To overcome this challenge, treatment should be closely linked to testing, integrated into primary care and made widely available in a cohesive model where patients can access both at a single-entry point.^8^

Test and treat strategies have been shown to be effective in HIV,^9^ Hepatitis C,^10^ human papillomavirus^11^ and sexually transmitted infections in Africa.^12^ For COVID-19 infection, modelling data demonstrates that for test and treat strategies to be successful in reducing the burden of severe cases, testing rates need to substantially increase in areas with suspected high positivity rates.^13^ Antigen-based testing has been shown to facilitate rapid expansion of testing, resulting in increased case identification and reduced test processing times, suggesting that antigen tests, rather than PCR-based tests, were essential to COVID-19 test and treat programs.^14^

Operational research, including qualitative research, is key to understanding context-specific insights and lessons in the early stages of program implementation.^15^ Previous research has explored the perspectives of healthcare workers on test and treat programs for HIV and malaria, highlighting the need to accommodate the increased workload from test and treat programs, ensuring the availability of testing and treatment commodities and the continuous need for healthcare worker training and refreshers.^16,17^ While some of these lessons learned may be transferable to the COVID-19 test and treat context, to date, operational research on COVID-19 test and treat program implementation is lacking.

This manuscript aims to understand the perspectives of key stakeholders, Ministry of Health, public sector personnel and healthcare workers on the early implementation of national COVID-19 test and treat programs in Ghana, Malawi, Rwanda and Zambia. This study is among the first to assess the barriers and enablers of a COVID-19 test and treat program from the perspectives of Ministry of Health/public sector personnel and healthcare workers in diverse LMIC settings.

## MATERIALS AND METHODS

### Study Setting and Site Selection

From late 2022 to 2023, nirmatrelvir/ritonavir was provided as standard of care in seven LMICs in sub-Saharan Africa (Ghana, Laos, Malawi, Nigeria, Rwanda, Uganda and Zambia), in partnership with the COVID Treatment QuickStart Consortium.^18^ In four of these countries (Ghana, Malawi, Rwanda and Zambia) nirmatrelvir/ritonavir was deployed to treatment sites as part of a multiple methods implementation research study in a test and treat program for treatment of mild-moderate COVID-19. Treatment site activation took place in each country at a different time, depending on drug availability and local regulatory approval: Ghana and Malawi, March 2023; Rwanda, February 2023; Zambia, December 2022. SARS-CoV-2 testing was combined with counselling and access to treatment for eligible patients. We conducted a multiple methods implementation research study divided into three components to provide simultaneous program learnings and improvements with program rollout. The study had three key operational research objectives: 1) program learnings, monitoring and evaluation; 2) patient-level program impact; and 3) key stakeholder perspectives.^18^ In this manuscript we explore the third research objective of understanding the perspectives of key stakeholders, including Ministry of Health (MoH), public sector personnel and health care workers on recommendations for improvement and strengthening of national COVID-19 T&T programs. Patient perspectives on recommendations for improvement and strengthening of the COVID-19 T&T program are presented elsewhere.

We used purposeful sampling to generate a list of study sites from the treatment sites across each country (**Fig 1**). Test and treat sites were primarily public health hospitals providing COVID-19 testing services (at the discretion of each country). Sites were selected in collaboration with the Ministry of Health in each country based on diversity within the sample, for example, rural and urban sites, geographic representation, caseload, and representation of different patient populations.^18^ All treatment sites followed established country protocols for COVID-19 patient testing and treatment with oral antivirals based on international guidelines, as described elsewhere.^19^

**Fig 1:** Map of Ministry of Health/public sector personnel and Healthcare Worker Interview locations.

### Participants and Sampling

We conducted in-depth, semi-structured interviews with individuals from the following stakeholder groups involved in the COVID-19 T&T program in each country: 1) MoH public sector personnel involved in developing and/or implementation of COVID-19 T&T policy and, 2) healthcare workers (HCWs) involved in administering COVID-19 testing and/or treatment at study sites.

MoH/public sector personnel were interviewed within each country to identify and select participants with a diversity of perspectives and experiences, and familiarity with the COVID-19 T&T program. HCWs were drawn from study sites through convenience sampling. HCWs were included if involved in administering COVID-19 testing or treatment at their site. Participants were free to decline participation without providing a reason.

Sample size was driven by information power.^20^ Based on previous work, our target sample size for the number of completed interviews was ten to 12 per stakeholder group, per country.^21^ Prior research suggests that a median of 8 to 16 in-depth interviews is needed to reach 80% and 90% saturation, respectively.^22^

### Study design

We conducted a descriptive qualitative study to explore recommendations for improvement and strengthening of national COVID-19 T&T programs.^23^ The Consolidated Framework for Implementation Research (CFIR) was used to guide qualitative data collection and analysis.

The CFIR is a comprehensive, meta-theoretical implementation science framework used to describe heterogeneity in implementation across settings, as well as the relative effect of key determinants in influencing implementation outcomes.^24^ The CFIR has been widely used to examine barriers and facilitators to program implementation and is the structural framework used to guide the qualitative aspects of this study. The CFIR contains five domains: intervention characteristics, inner setting, outer setting, individual characteristics, and implementation process. We used CFIR to guide our interview guide development, data collection, and analysis.

Semi-structured interview guides were developed for each stakeholder group using *a priori* selected CFIR domains and constructs of:

- *Implementation process (planning and reflecting and evaluating): activities and strategies used to implement the test and treat program*
- *Intervention characteristics (intervention complexity and adaptability): complexity and adaptability of the test and treat program that may impact implementation*
- *Inner setting (relative priority): the prioritization of the test and treat program compared to other healthcare programs or initiatives*

MoH/public sector personnel interview guides addressed policy considerations for implementation of the program (**Supplemental 1**) and healthcare worker interview guides addressed facility-based barriers and challenges in implementation of the program (**Supplemental 1**).

### Data Collection

An experienced team of qualitative researchers from Duke University, supported by Clinton Health Access Initiative staff, trained country-level data collectors on interviewing skills, note-taking skills, audio recording and obtaining informed consent. Written or verbal informed consent was obtained prior to administering all interviews. The interview duration was 30-45 minutes and English was the predominant language used to conduct the interviews across all stakeholder groups in all countries, except Rwanda, which was primarily in Kinyarwanda. All interviews for HCWs were conducted in-person. Some interviews for MoH and public sector personnel were conducted via Zoom or phone when in-person meetings were not feasible.

Interview questions were open-ended, and probes (“Can you tell me more?” “Can you give me an example?”) were used as needed to elicit more detailed information or to clarify interviewee responses. Most interview sessions included an interviewer and note-taker. Where two people were not available, the interviewer conducted the session and returned to the audio recording to complete the note-taking form. Demographic details of interviewees were obtained at the beginning of the interview. Structured note-taking forms were used to document key points both during and immediately after the interviews, allowing for real-time reflections and additional observations. The interviews were audio-recorded using a device-based recording app, uploaded and saved through SurveyCTO® as an additional resource for both data collectors and qualitative researchers. Audio recordings of interviews were available to the data collection staff to ensure accurate completion of the note-taking forms. Where interviews were conducted in languages other than English, data collectors translated responses and completed the note-taking forms in English. Only audio interviews conducted in English were available to the qualitative research analysts to obtain verbatim quotes. Demographic details, note-taking forms and audio interviews were uploaded to a secure, encrypted server for analysis.

### Analysis

Qualitative interview data was analyzed with a rapid content analysis technique,^25,26^ using Microsoft Excel version 2019 to support coding and analysis. We focused our analysis on the *Implementation Characteristics* domain and specifically the *Intervention Adaptability* construct to inform timely program planning, tailoring, and evaluation.^27^ An experienced pair of qualitative researchers (SG and SN; JM and CM) reviewed all note-taking forms for each stakeholder group to develop interview debrief notes using thematic analysis and the matrix method, guided by CFIR.^28,29^ Rigor and validity were established by independently coding and summarizing the data, discussing emerging codes and thematic groupings during meetings, and receiving input from a third qualitative researcher (MO) to resolve any discrepancies.

### Ethics

This study was approved by the Duke University Institutional Review Board (Pro00111388), The study was also approved by local institutional review boards in each country: the Ghana Health Services Ethics Review Committee (017/11/22), the National Health Sciences Research Committee, Malawi (#23/03/4025), the Rwanda National Research Ethics Committee (105/RNEC/2023) and the Institutional Review Board ERES Converge (Zambia) (23-Jan-023).

## RESULTS

### Participant Characteristics

**Table 1** shows the demographic information for the Ministry of Health/public sector personnel and healthcare worker participants (stakeholders) in the study, including the data collection period, roles of participants and languages the interviews were conducted in. Ten to 14 interviews were conducted per stakeholder group in each country with a total of 97 interviews across all countries. Interviews were conducted between 01/03/2023 – 01/06/2024. The data collection period differed slightly in each country due to varying introduction times of nirmatrelvir/ritonavir and subsequently program start in each country. Interview recruitment was commenced within three months of the official launch of the test and treat program in each country. Where verbatim quotes were able to be obtained, these have been presented as exemplars of identified constructs/themes. Interviewer notes are presented where interviews were conducted in a language other than English and verbatim quotes could not be obtained.

**Table 1:** Demographics of study participants.

|  |  | Ghana |  | Malawi |  | Rwanda |  | Zambia |  |
| --- | --- | --- | --- | --- | --- | --- | --- | --- | --- |
| Data collection period |  | August 2023 - March 2024 |  | July - December 2023 |  | July 2023 |  | April - December 2023 |  |
| Participant role |  | MoH | HCW | MoH | HCW | MoH | HCW | MoH | HCW |
| Number of study participants |  | 10 | 13 | 13 | 12 | 10 | 14 | 13 | 12 |
| Gender | Male | 70% | 92% | 46% | 67% | 20% | 43% | 46% | 75% |
|  | Female | 30% | 8% | 54% | 33% | 80% | 57% | 54% | 33% |
| Age range | Younger than 35 | 0% | 62% | 54% | 50% | 50% | 64% | 31% | 75% |
|  | 35 or older | 100% | 38% | 46% | 50% | 50% | 36% | 69% | 25% |
| Data collection language (majority language vs all languages, MOH/HCW) |  | English | English | English | English | Kinyarwanda and English | Kinyarwanda | English | English |

**Figure 1** demonstrates the location where interviews were conducted with participants. These locations include a mixture of urban and regional centers where the T&T program was operating.

### Interview Themes

Four major themes of recommendations to strengthen COVID-19 T&T programs were identified in the interviews that were consistent across all groups and countries (**Table 2**). In addition, one further theme was identified that was specific to the HCW participants in all countries (**Table 2**).

**Table 2:**
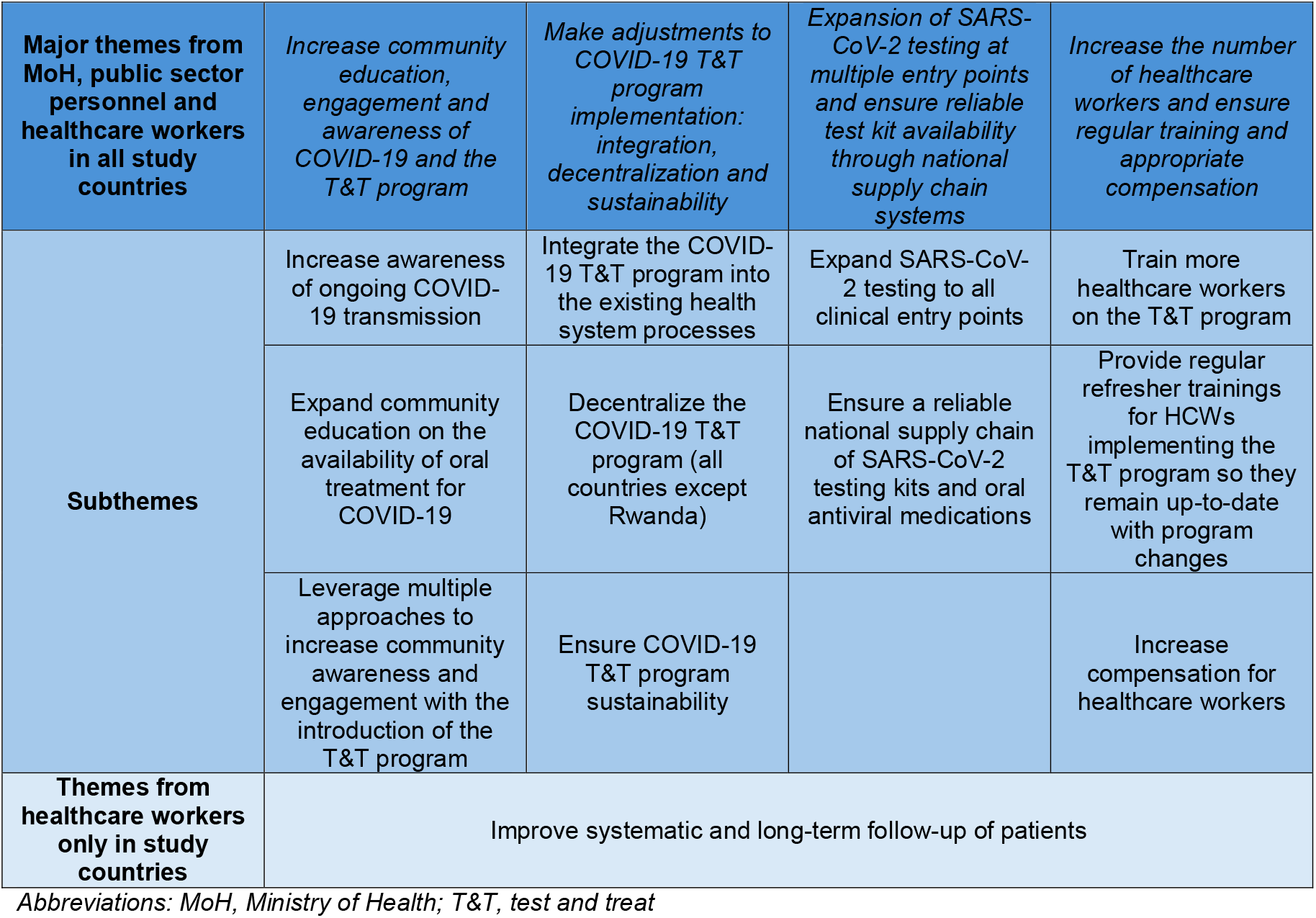
Summary of findings based on CFIR Construct.

#### 1. Increase community education, engagement and awareness of COVID-19 and the T&T program

This theme describes awareness of ongoing COVID-19 transmission in the community and the importance of community education on the availability of oral treatment for COVID-19. This theme also describes leveraging multiple approaches to increase community engagement with the COVID-19 T&T program.

##### Increase awareness of ongoing COVID-19 transmission

MoH personnel and HCWs underscored the necessity of emphasizing ongoing SARS-CoV-2 transmission which was waning due to COVID-19 pandemic fatigue. Stakeholders also highlighted the value of community education on COVID-19 infection to combat stigma related to the illness and the subsequent minimization of symptoms in the community.

> *“…people [in the community] still believe there is no COVID, it’s something else. Because of that type of stigma, even acceptance on these drugs, even on the test…for the rural areas, without community sensitization, we will have a very big challenge*.*”* MoH, Malawi, July 2023.

##### Expand community education on the availability of oral treatment for COVID-19

MoH personnel and HCWs reported that increased public knowledge was needed, particularly among the elderly and in rural areas, that effective oral treatment was available for mild-moderate COVID-19 infection.

> *“As of now, the majority of people within our catchment area still do not know that there is even a treatment for COVID-19, in terms of the mild to moderate disease…I think there is [a need for] massive education*.*”* HCW, Ghana, August 2023.

Increased knowledge of oral treatments available for COVID-19 may encourage community members to seek medical care, instead of avoiding healthcare facilities due to concerns around the need for isolation or quarantine.

> *[interviewer notes from HCW, Rwanda, July 2023] It would be important to educate and communicate to the patient or the population in general about this [nirmatrelvir/ritonavir] treatment. Because it would be easier for the patient to come to the hospital if they know there is treatment, and they are going home with it. It will remove the social stigma of thinking that [they are] going to be put in quarantines and isolation*.

Local engagement was also suggested as key to encouraging community members to accept free treatment for COVID-19, and address some public concerns about free medicines being in a trial phase.

> *[interviewer notes from HCW, Rwanda, July 2023] Most patients that we meet here have no clue about this medication. So, when you prescribe the drug and tell them that its free, they become skeptical to take it, there is a stigma around free drugs here, patients think it’s not good or maybe it’s still in trial phase. But I think if we can talk about this medication beforehand, this would really help … us the health workers*.

##### Leverage multiple approaches to increase community awareness and engagement with the introduction of the T&T program

Increasing community awareness of the T&T program through multi-sectoral community engagement was emphasized by stakeholders. Various approaches were suggested, such as through community and spiritual leaders, media (television, radio, podcasts, pamphlets, posters, billboards), community campaigns, health talks, megaphones in public, promotional material such as t-shirts or using large events and fairs through the Ministry of Health.

> “*You need to look at the aspect of community engagement, very important – a multi-sectoral approach. You have to talk to the community. Use different forms of mass media - radio, TV, podcasts, whatever is available, to get the information out there. Because right now most people do not know about [COVID-19 treatment]*.*”* HCW, Zambia, December, 2023.

It was also suggested that raising community awareness could help generate demand for SARS-CoV-2 testing and treatment, encouraging individuals with symptoms to get tested and seek care at a clinic within the recommended timeframe.

> *If we could get large-scale form of advertisement or education forum on the radio stations and television, something that runs for a while before the program launches, that would really help*.*”* HCW, Ghana, August 2023.

#### 2. Make adjustments to COVID-19 T&T program implementation: integration, decentralization and sustainability

This theme outlines the importance of integrating the COVID-19 T&T program into existing health systems, decentralizing the program to lower-level facilities and ensuring program sustainability.

##### Integrate the COVID-19 T&T program into the existing health system processes

MoH personnel and HCWs recommended that the COVID-19 T&T program should be integrated with other similar and existing services and programs. Suggestions for program integration included testing for SARS-CoV-2 for all those presenting with respiratory symptoms at emergency triage, or testing for SARS-CoV-2 in existing tuberculosis clinics, and offering treatment to those who are eligible.

> “…*it would be important to find ways to integrate the test and treat program in healthcare services like other infectious diseases. So, when people come with respiratory symptoms, we should be able to exclude COVID-19 at triage while screening*.” HCW, Zambia, December, 2023.

##### Decentralize the COVID-19 T&T program

The COVID-19 T&T program should be decentralized from the central hospitals (tertiary and secondary facilities) to smaller hospitals and local health care centers (primary care facilities) as primary care level facilities are where most people access their health care. Countries should also consider options beyond central health facilities and allow for community distribution points, such as local pharmacies, to increase access to testing. Decentralization of the program would support COVID-19 treatment at the location where a patient tests positive for SARS-CoV-2, and would improve early case detection.

> *“There should be multiple distribution points in the public…so that there is increased access. So it needs to have several distribution points [in the] community, schools*…*just everywhere*.*”* MoH, Zambia, October 2023.

Program testing in central hospitals should also be decentralized to clinics where people are being seen, as some patients who travel to the laboratory for SARS-CoV-2 testing do not return to the clinic to obtain their test result and receive treatment.

> “*For us, sometimes we tend to lose patients [with COVID-19] in transit, maybe they are going to the lab to be tested for COVID-19… then they tend to let go of that patient because they don’t go back to the technician. So, maybe for us not to lose patients…we try to decentralize the testing and make it available at the place [the clinic] where people are being seen*.*”* HCW, Malawi, September 2023.

This sub-theme did not apply to Rwanda where the COVID-19 T&T program was implemented nationwide through a decentralized health system.

##### Ensure COVID-19 T&T program sustainability

It is important to ensure that the COVID-19 T&T program is cost-effective and sustainable for the government to continue after program initiation. Support from all stakeholders involved, especially senior management, is also needed for program sustainability.

> *“When programs start, when they’re donor-aided, the donors are putting in a lot of resources. When they [donors] leave for the country to take over the program, and it’s too expensive, programs tend to die down. Because the priorities are different and institutions may not have resources to continue the program. So something that can be sustained, even with limited resources, I think that would be great*.*”* MoH, Malawi, September 2023.

Stakeholders emphasized that the COVID-19 T&T program should not be a siloed or stand-alone program, but instead it should be integrated into existing processes and systems to remain sustainable.

> *“The assessment and the treatment has to be done where outpatient interventions are being done…the intervention has to be within the system of the facilities and not a parallel system…if it has its own parallel system, once the resources are gone, government will not continue [the program]. But if it is embedded within the current system in the OPD (outpatient department) setup then it’s fine*.*”* MoH, Malawi, July 2023.

#### 3. Expansion of SARS-CoV-2 testing at multiple entry points and ensuring reliable test kit availability through national supply chain systems is needed for program scale-up

This theme explores the expansion of SARS-CoV-2 testing at all clinical entry points to care and ensuring a reliable supply chain of testing kits and treatment for COVID-19.

##### Expand SARS-CoV-2 testing to all clinical entry points

SARS-CoV-2 testing should be expanded to all facilities and hospital entry points, including non-inpatient locations i.e. HIV clinics, non-communicable diseases clinics and maternal child health clinics.

> *“Beyond just allowing the patients to get tested in inpatient facilities, we expanded the testing. We encourage testing to be done in HIV clinics, so even when patients go for their routine visits, they can be able to get tested [for COVID-19]. I think one of our other strategies is to encourage [testing] in noncommunicable disease clinics, if you’re going for a diabetes testing you can get a COVID test. We also want to work with the maternal child health clinics because those are congregant settings and people come whether or not they just have flu-like symptoms*.*”* MoH, Zambia, May 2023.

Testing for SARS-CoV-2 should also be promoted among all patients with flu-like symptoms, rather than just those suspected of having a COVID-19 infection, to increase access to COVID-19 oral treatment for eligible patients.

> *“…for the full implementation of the test and treat [program] maybe the clinicians, they have to, as much as possible, any client or patient who is suspected [of having COVID-19] has to be screened. So they have to scale up [testing and treating]…by testing as much as possible*.*”* MoH, Malawi, September 2023.

##### Ensure a reliable national supply chain of SARS-CoV-2 testing kits and oral antiviral medications

SARS-CoV-2 testing should be available at all possible health facilities. This is dependent on a reliable and timely supply chain at all levels of the health system and integration of the testing kit supply chain with existing systems.

> *“… to ensure that … [the COVID-19 T&T] program continues and we see it being successful, it means that other things have to be in place in order for it to work. The most important being normalizing or making sure that the supply chain or central commodity is working, so the drugs, the testing kits, should always be available…”* HCW, Zambia, July 2023.

Oral antiviral medications should also be reliably obtained at locations where SARS-CoV-2 testing is available, so community members are able to obtain testing and treatment at the same location.

> “*In terms of commodities…we have to think about the fact that if we open this up to all facilities…we have to think about ensuring that we have the antivirals always available to be able to give to people and also test kits as well*.*”* MoH, Ghana, November 2023.

#### 4. Importance of increasing the number of HCWs and ensuring regular HCW training and appropriate compensation

This theme describes the importance of training more HCWs on the T&T program, ensuring HCWs are kept up-to-date on program changes and increasing compensation for HCWs.

##### More HCWs need to be trained on the T&T program

HCWs at all levels and disciplines should be trained on the T&T program, so that more patients can be reached. Healthcare workers trained through the train-the-trainer model should also possess a thorough understanding of the program and its protocols to ensure it is implemented properly.

> *“…we need a serious training on health workers…especially those that are on the ground, those are the ones that need to really know. Because sometimes…you find you only invite the big bosses, and when they are done they will not disseminate the information…to those that are doing the skill on [the] ground*.*”* HCW, Zambia, Interview conducted in July, 2023.

Increasing the number of HCWs trained on the T&T program provides a buffer for the program if staff are absent (i.e. due to vacations or departure from the facility).

> *“Even when I’m not around, doesn’t mean the work shouldn’t go on, so somebody should have stepped in. But because we are understaffed and you are not around [core trained staff] it means there is a gap…you get people, you train them – the next day they are leaving. There is a big gap*.*”* HCW, Ghana, December 2023.

##### Regular refresher trainings for HCWs implementing the T&T program are crucial to remaining up-to-date with program changes

Training on the T&T program should be repeated for HCWs through ‘refreshers’ and ‘updates’ to ensure that HCWs are aware of changes in COVID-19 clinical guidelines, and they are motivated and encouraged to maintain program implementation at their facilities. Refresher training is also an avenue for ongoing mentorship of staff who are administering the program at smaller hospitals and health posts.

> *“In the medical field, we need to make sure we are doing the right things. So if you’re not comfortable doing [a COVID-19 T&T program task] and I don’t think anyone can force you. And it can be like ‘it’s been really long since we did this testing, we need a refresher*.*’”* HCW, Malawi, September 2023.

##### Increase compensation for HCWs

Increasing compensation for HCWs on the T&T program would offset out-of-pocket expenses such as phone subscription plans and transportation fees incurred by HCWs in program implementation. Increased compensation or incentives may also be necessary to compensate for the increased workload of the COVID-19 T&T program on top of existing clinical duties, or to help prioritize the program and maintain staff interest in program activities.

> *“If this program is to succeed on a national level, let incentives be placed, especially for people in the peripheries [rural/remote areas] – because in the peripheries, staffing is poor…you’ll find someone is on call every other day and on top of that they are a COVID focal point person. That means after they are on call, they have to collect the [program] data*.*”* HCW, Zambia, July 2023.

### Additional theme from healthcare workers

#### Improvement of patient follow-up

More systematic, longer-term and in-person follow-up of patients treated with oral antivirals for COVID-19 is needed to monitor for medication adherence and identify drug side-effects.

> *[interviewer notes from HCW, Rwanda, July 2023] [follow-up was conducted via telephone only] Our main operational challenge to date is in the follow up process. We cannot be sure the patients are really taking the medications when they reach home, moreover the fact that we are not able to go and follow them up at home in person, ask them about their progress, ask about the taste of the drug and counsel if they have difficulties adhering to the drugs is the main challenge we face*.

HCWs suggested that follow-up may be easier for patients if transportation was provided for follow-up visits, or if phone credit was provided to patients to be contacted for follow-up.

> *“I also foresee the challenge of doing the [patient] follow ups on a national level. It’s not every facility that will allow, or that will looking at the cost involved, would put staff in a vehicle and say that ‘go and make a follow up to this person in this community, in this community’ or even give credit for that*…*credit as in making the calls, airtime. It’s not every facility that will do that. At the national level, the dynamics would be different*.*”* HCW, Ghana, September 2023

## DISCUSSION

This manuscript describes four key themes identified by key stakeholders (MoH and public sector personnel and HCWs) in scaling up COVID-19 T&T programs: 1) increasing community education, engagement and awareness of COVID-19 and the T&T program, 2) adjusting the T&T program to ensure program integration, decentralization and sustainability, 3) expanding SARS-COV-2 testing and ensuring availability of testing kits and oral antivirals through reliable national supply chains, and 4) ensuring ongoing training and support for HCWs on the COVID-19 T&T program. We found that countries were largely united in their recommendations, suggesting that a unifying framework for introducing new drugs through T&T programs during future public health emergencies in Sub-Saharan countries may be possible.

A key finding of this study was that major recommendations from key stakeholders in program implementation were consistent across all four countries involved. These recommendations transcended significant differences in economy, culture, health system structure, timing of availability of nirmatrelvir/ritonavir and local COVID-19 infection and vaccination rates. To our knowledge, the consistency of recommendations across countries implementing test and treat programs has not been described. Our study’s flexible protocol, which allowed for minor adjustments in drug eligibility (based on national guidelines), adaptations to existing health system structures (i.e., centralized vs. decentralized systems) and region-specific approaches to program promotion, were crucial in enabling countries to implement locally tailored programs.

The convergence of recommendations aimed at enhancing program feasibility and acceptability suggests the potential to develop a shared “roadmap” for introducing new drugs through T&T programs during future pandemics. Such a framework could be valuable for pandemic preparedness in the four countries that participated in this study and may also be applicable to other Sub-Saharan African nations or low- and middle-income countries, particularly when implementation strategies can be customized to fit local contexts.

Community engagement and awareness in scaling up COVID-19 T&T programs emerged as a key lesson learned in review of the program in the first 12 months. Stakeholders identified two key aspects of community engagement requiring greater focus. First, increase awareness of ongoing COVID-19 transmission in the community, including understanding who was at risk for severe disease. The decrease in risk perception was particularly notable in countries where program implementation and interviews took place following the World Health Organization’s May 5, 2023 announcement declaring an end to COVID-19 as a Public Health Emergency of International Concern.^30^ Additionally, stakeholders noted the persistence of stigma surrounding COVID-19 testing and diagnosis which discouraged individuals from seeking care when symptomatic. This stigma was often likened to the stigma associated with HIV/AIDS, with similar negative impacts on individuals’ willingness to seek care or disclose their COVID-19 status.^31,32^

The second aspect of community engagement and awareness highlighted in the study was community recognition that a new oral treatment was available for COVID-19. The marketing and educational activities aimed at creating awareness, interest and demand for nirmatrelvir/ritonavir were different in each country. The local demand for treatment depended on the timing of drug introduction with local COVID-19 incidence, with lower demand for the drug in settings with fewer cases. Local COVID-19 incidence trends also impacted COVID-19 vaccine uptake,^33^ and other local public health concerns. In Zambia, nirmatrelvir/ritonavir was introduced during a wave of COVID-19 infection with widespread government promotion and support, increasing the demand for and awareness of the product. In contrast, the new drug was introduced in Malawi at a time when SARS-CoV-2 incidence was low and another emergency, Tropical Cyclone Freddy,^34^ was taking place, which impacted the early uptake of the drug.

To our knowledge, this is the first study examining implementation of an oral antiviral treatment for COVID-19 through a test and treat program in Sub-Saharan Africa. While comparison with similar studies is not possible, our study results are supported by COVID-19 vaccine study findings from the region demonstrating that community awareness — communication, mobilization and education — is a crucial aspect of vaccine uptake.^35,36^ Vaccines for COVID-19, unlike other mass vaccination campaigns, were rapidly rolled out, with limited information available related to long-term safety data and duration of protection. In the pandemic setting, community awareness campaigns were conducted quickly and selectively, with less social listening and monitoring of public perceptions to guide demand-generation efforts.^37^ The introduction of oral antivirals for COVID-19 in the countries studied faced similar challenges to those encountered with vaccines, in that there was an urgent need to rapidly increase access. While community awareness efforts can be both time-intensive and costly, our findings support the centrality of these efforts to demand-generation for new antiviral introduction a public health emergency.

The importance of ongoing sustainability and decentralization of the T&T program emerged as the second key theme. Stakeholders emphasized the importance of integration of the COVID-19 program within existing health services rather than creating vertical or siloed programs. Integration of the COVID-19 T&T program has already commenced in some countries participating in this study, particularly within existing national tuberculosis (TB) programs. A model of reciprocal testing for COVID-19 and TB programs where testing and follow-up for both diseases are performed at a single entry-point for patients presenting with respiratory symptoms. TB prevention efforts decreased globally at the onset of the COVID-19 pandemic,^38^ and integration of COVID-19 and TB services has been identified, and successfully implemented, to address the challenges in resource allocation during the pandemic.^39,40^

The decentralization of the COVID-19 T&T program was also identified as essential to its sustainability, emphasizing the need to provide SARS-CoV-2 testing and treatment at local community health facilities and pharmacies. A review of oral antiviral implementation for COVID-19 in Sub-Saharan Africa suggested that deploying the treatment through nurse-led, doctor-supported primary care facilities is the optimal model, as it offers a feasible, scalable, and sustainable approach with the greatest potential impact on health.^41^ Studies from Asia have also shown that a key factor in oral antiviral uptake has been the recommendation of COVID-19 treatment from a primary care physician.^42,43^ Task-shifting among community health service personnel, similar to HIV and Hepatitis C programs, will be important to any successful program to decentralize COVID-19 testing and treatment.^44,45^ Delegation of tasks to healthcare workers who do not require advanced medical training may allow primary health services to implement the COVID-19 T&T program, improving patient access to antivirals within the window period and health-system efficiency.

The third theme emerging from this study was the need for ongoing training, education, and improved renumeration for healthcare workers. Ongoing training and education on the COVID-19 T&T program was emphasized as essential to program feasibility. The training challenges raised by healthcare workers were 1) orienting new hires, 2) updating training as guidelines change and 3) the efficacy of the ‘train-the-trainer’ model and the success of the cascade of training. These hurdles have been overcome in similar programs with the provision of job aids, standardized manuals, refresher trainings and telehealth to deliver training courses, clinical mentoring and new guideline dissemination.^46,47^

Healthcare workers also raised the challenges of implementing the COVID-19 T&T program as an extra clinical duty above their existing responsibilities. Countries in our study were not able to increase renumeration for healthcare workers implementing the COVID-19 T&T program due to significant other expenditures. The additional workload for healthcare workers came amid a decrease in income for some during the COVID-19 pandemic, challenges with access to personal protective equipment, an increased risk of COVID-19 infection and mortality, and burnout from the demands of their occupation.^48-50^ Future test and treat programs should consider whether increased renumeration and support should be provided to healthcare workers undertaking programmatic duties in addition to their regular clinical duties.

### Limitations

A limitation of this study is that the COVID-19 T&T program was implemented after the peak in SARS-CoV-2 incidence in most of the implementing countries, and in large part after May 2023 when <u>World Health Organization</u> declared COVID-19 was no longer an emergency. If the program were implemented early in the pandemic, implementation findings may have been different. Additionally, data were collected during the early phase of implementing the program. The program was in its first year of operation when interviews were conducted, which likely influenced the strong emphasis stakeholders placed on community awareness as a critical component for program scale-up. At that early stage, community awareness efforts in some countries may not reflect the current level of awareness as the program has matured. However, the goal of the operational research approach was to provide timely findings to local program implementers for improvement and scale-up, making stakeholder perspectives from the program’s initial months essential. Future operational research studies focused on rapid learnings from implementation could benefit from conducting stakeholder interviews both early and later in the program timeline to assess whether community awareness improves as the program progresses.

A second limitation is that the qualitative data analysts did not have access to verbatim transcripts in English (their spoken/written language). To ensure rapid results for program implementation, we conducted interviews in the language most comfortable for both the interviewee and interviewer and did not transcribe or translate the audio files. As a result, we relied on interviewer notes rather than transcripts to capture the interviewee’s perspective. Additionally, interviews conducted in a language other than English may have lost some of the nuance of the interview during translation. Future research could benefit from transcribing and formally translating interviews to allow for the inclusion of direct quotations from all stakeholders, regardless of the language spoken, albeit at a time and resource cost consideration.

Finally, response bias could have been a limitation of this study if participants provided answers that were more aligned with what they thought was expected or desired, rather than expressing their true feelings or experiences. To mitigate this bias, we used data collectors who were not involved in the program’s design or implementation, with the aim of ensuring they were perceived as impartial outsiders with no vested interest in the program.

## CONCLUSION

This study examines the introduction of a novel antiviral treatment, nirmatrelvir/ritonavir for COVID-19, through a test and treat program in four African countries. Qualitative results from the first twelve months of program implementation identified actionable changes common to all four countries for improved program feasibility, acceptability and scale-up, with a focus on community education, program integration, decentralized service delivery and ongoing healthcare worker training. Our findings suggest that a unifying framework for introducing new drugs through T&T programs during future pandemics in the four countries that participated in this study, or other Sub-Saharan countries may be possible, especially when implementation strategies can be customized to fit local contexts.

## Data Availability

NA

## Acknowledgements

We acknowledge government partners and healthcare workers in QuickStart program countries for their dedication in implementing this program. We recognize Americares for their collaboration in this consortium as partners involved in the logistics and delivery of donated products. This work was funded by Pfizer, the Open Society Foundations, and the Conrad N. Hilton Foundation. The donors have no role in the design or analysis of this implementation research study.

We acknowledge all members of the COVID Treatment QuickStart Consortium:

Opeyemi Abudiore, Chukwuemeka Agwuocha, Oluwatosin Akinwande, Okechukwu Amako, Maame Nkansaa Asamoa-Amoakohene, Jason Blevins, Vishal Brijlal, Ellen (Nellie) Bristol, Phyllis Chituku, Maria Corcorran, Khamsay Detleuxay, Anthony Bless Dogbedo, Somsy Dongpadith, Devan Dumas, Gary Edson, Leslie Emegbuonye, Fiona Gambanga, Michelle Gao, Maria Grau-Sepulveda, Prudence Haimbe, Elina Urli Hodges, Lorraine Kabunga, Joseph Kalua, Pax Axell Karamage, Davis Karambi, Brenda Kateera, Bounxou Keohavong, Philip Kimani, Robert Kirungi, Megan Knox, Monica Leyva, Viengsakhone Louangpradith, Folu Lufadeju, Emily Macharia, Alexio Mangwiro, Wenhui Mao, Michael Merson, Jonathan Mtaula, Moses Mukiibi, Mwaba Mulenga, Lloyd Mulenga, Tamara Mwenifumbo, Vennie Nabitaka, Ciru Wanjiru Ndichu, Alida Ngwije, Ijeoma Okoli, Katharine Olson, Nere Otubu, Sompasong Phongphila, Christian Ramers, Simene Sangha, Hilda Shakwelele, Alan Staple, Jessica Tebor, Dyson Telela, Laine Thomas, Wesley Tomnos, Evarist Twinomujuni, Sabine Umuraza, Garrett Young

## Author Contributions

SN and BG contributed equally to this paper. Conception and design of the study: SN, BG, SG, CB, NH, JJ, EM, KU, CW, MO and HB. First draft of the manuscript: SN, BG, SG. Data analysis: SN, SG, CM, JM, MO. Critical revision of the manuscript for important intellectual content: All authors. All authors take responsibility for the accuracy of the discussion and had authority over manuscript preparation and the decision to submit the manuscript for publication.

