## Supplemental Information for "Recommendations for Enhancing COVID-19 Test and Treat Programs in Four African Countries: Insights and Strategies from a Qualitative Study"

**SUPPLEMENTAL MATERIAL**

**CONTENTS**

**Figure 1:** MoH/Public Sector Personnel Interview Guide	Page 2

**Figure 2:** Healthcare Worker Interview Guide	Page 3

**Figure 3:** Patient Interview Guide	Page 4

**Figure 1: MoH/Public Sector Personnel Interview Guide**

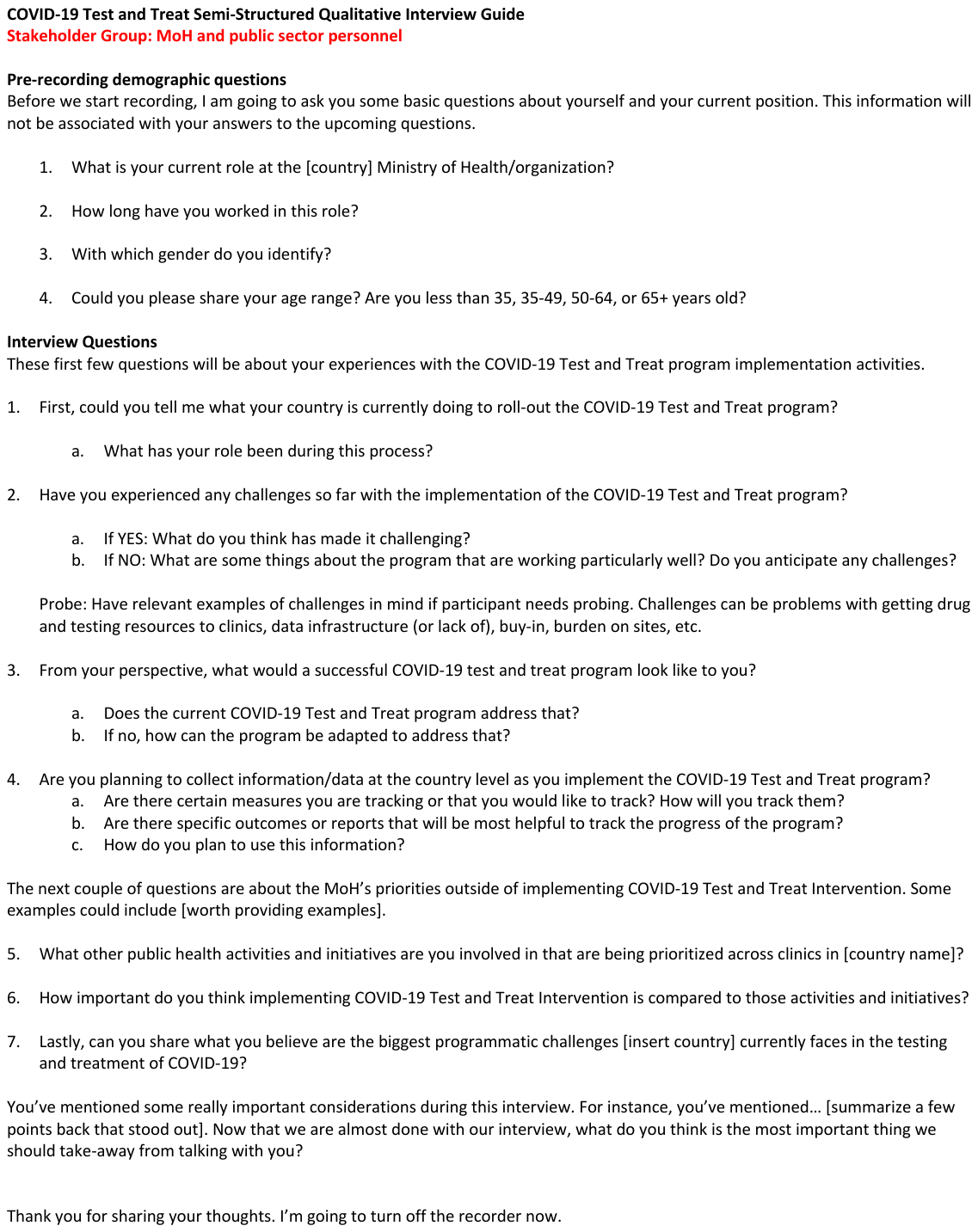

**Figure 2: Healthcare Worker Interview Guide**

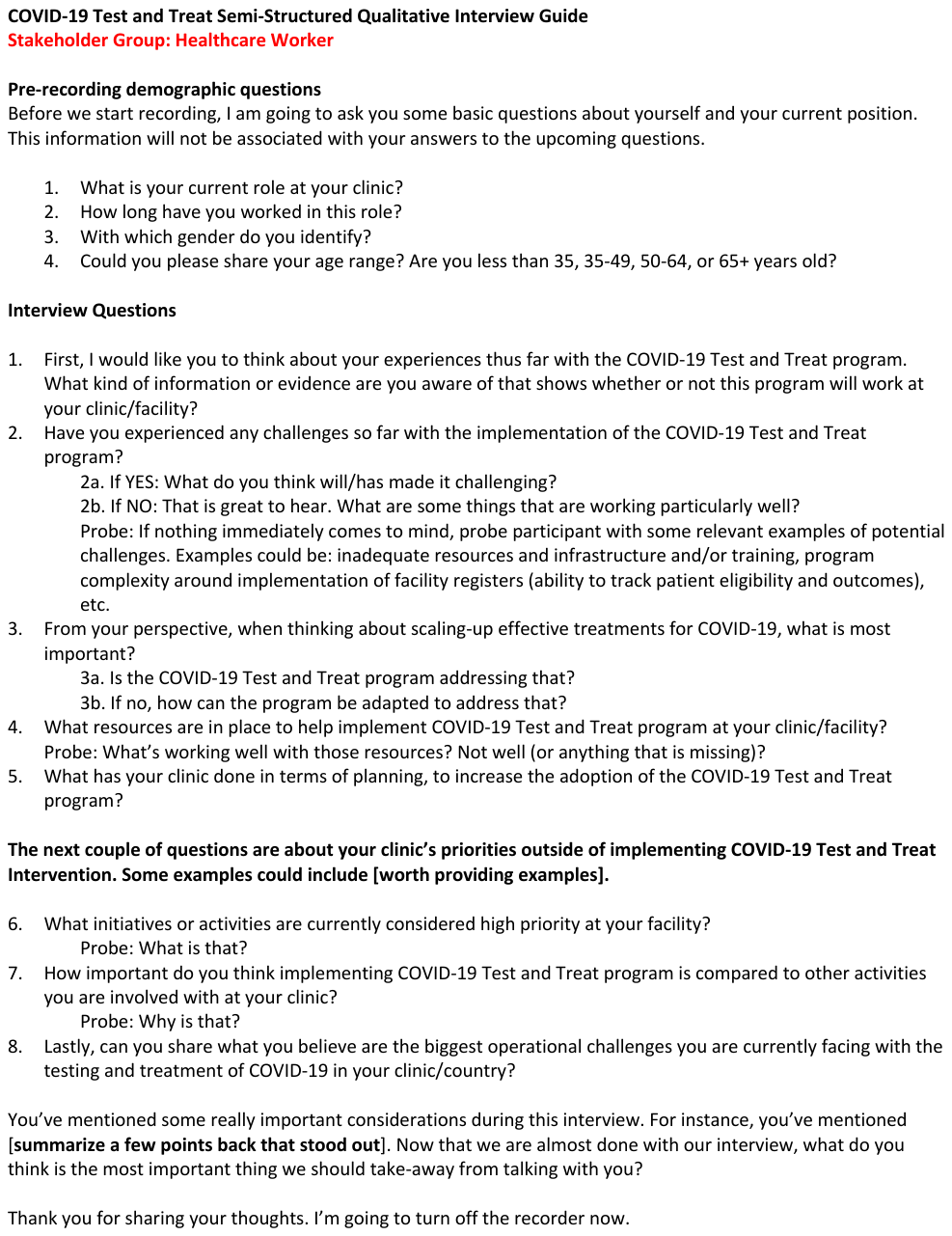

**Figure 3: Patient Interview Guide**

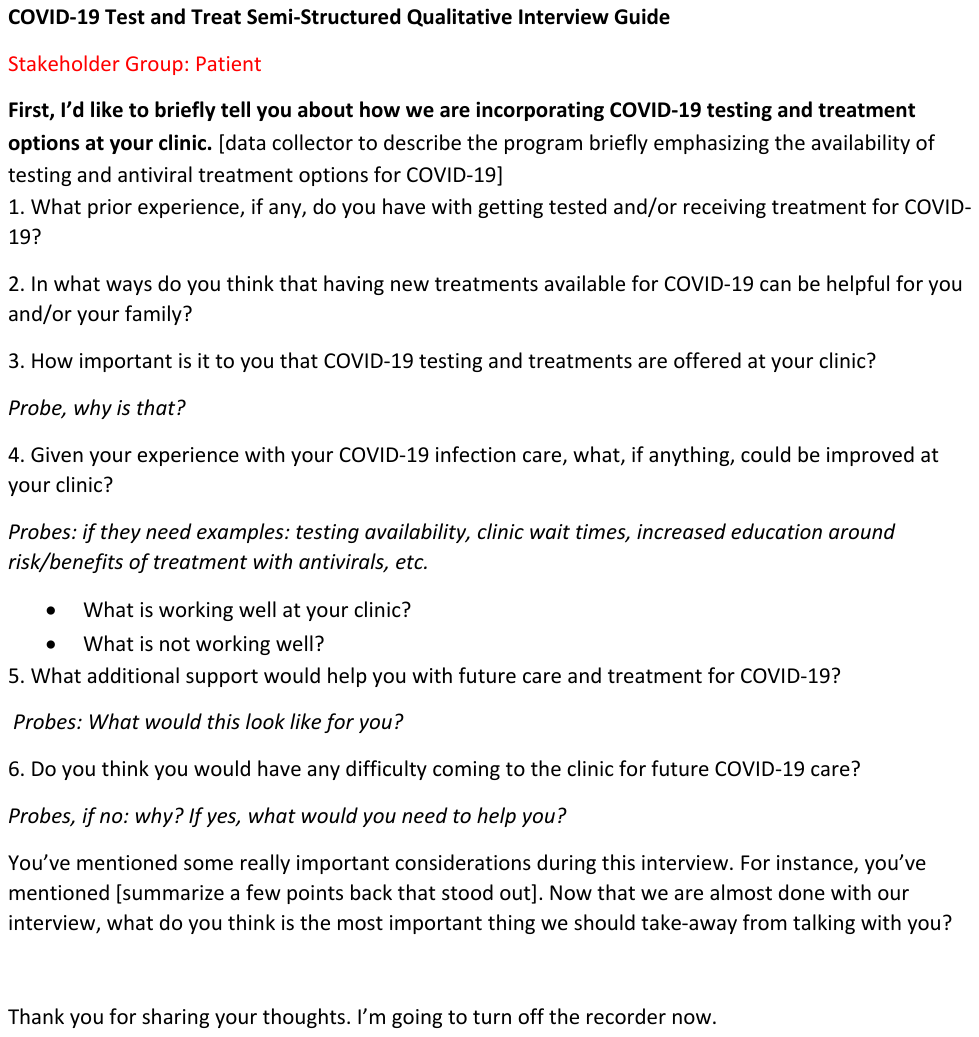
